# Acute hepatitis C infection among adults with HIV in the Netherlands: a capture-recapture analysis

**DOI:** 10.1101/19002097

**Authors:** Tamara Sonia Boender, Eline Op de Coul, Joop Arends, Maria Prins, Marc van der Valk, Jan T.M. van der Meer, Birgit van Benthem, Peter Reiss, Colette Smit

**Author notes:** Corresponding author T. Sonia Boender, PhD; /.

## Abstract

**Background:** Reliable surveillance systems are essential to assess the national response to eliminating hepatitis C virus (HCV), in the context of the global strategy towards eliminating viral hepatitis.

**Aim:** We aimed to assess the completeness of the two national registries of acute HCV infection in people with HIV, and estimated the number of acute HCV infections among adults with HIV in the Netherlands.

**Methods:** For 2003-2016, cases of HCV infection and reinfection among adults with a positive or unknown HIV-serostatus were identified in two national registries: the ATHENA cohort, and the National Registry for Notifiable Diseases. For 2013-2016, cases were linked, and two-way capture-recapture analysis was carried out.

**Results:** During 2013-2016, there were an estimated 282 (95%CI: 264-301) acute HCV infections among adults with HIV. The addition of cases with an unknown HIV-serostatus increased the matches (from N=104 to N=129), and a subsequently increased the estimated total: 330 (95%CI: 309-351). Underreporting was estimated at 14-20%.

**Conclusion:** In 2013-2016, up to 330 cases of acute HCV infection were estimated to have occurred among adults with HIV. National surveillance of acute HCV can be improved by increased notification of infections. Surveillance data should ideally include both acute and chronic HCV infections, and be able to distinguish between acute and chronic infections, and initial and reinfections.

**Classifications:** The Netherlands; sexually transmitted infections; hepatitis C; HIV infection; Surveillance; epidemiology

## Introduction

Hepatitis C virus (HCV) infections are generally uncommon in the Netherlands, with a chronic HCV prevalence of <0.2%[1]. However, an increase in the number of acute HCV infections and reinfections among men who have sex with men (MSM) who are HIV-positive has been reported since early 2000[2–4]. Detection, diagnosis and registration of acute HCV infections are crucial to measure trends in the epidemic, and plan appropriate public health and clinical interventions, such as prevention programs for those at risk, targeted testing, increasing treatment uptake, and contact tracing to reduce subsequent transmission. In addition, treating chronic HCV infections with direct acting antivirals (DAAs) is expected to strongly reduce, but not eliminate, the HCV epidemic among HIV-positive MSM[5,6]. Despite high DAA uptake in HIV/HCV co-infected people in the Netherlands[7], ongoing HCV transmission and reinfection continue to occur.

In the Netherlands, the Dutch HIV Monitoring Foundation (*Stichting HIV Monitoring, SHM*)[8] registers acute HCV infection among people with HIV since 2000, and the National Registry for Notifiable Diseases at the National Institute for Public Health and the Environment (*Rijksinstituut voor Volksgezondheid en Milieu, RIVM*)[4] registers acute HCV infections among people irrespective of HIV-serostatus since 2003. While the RIVM and SHM have reported similar trends of acute HCV infection over the years, consistently more cases were registered among HIV-positive MSM by the SHM, compared to the RIVM[4,9].

Reliable surveillance systems, monitoring and evaluation are essential to assess the national HCV response, in the context of the global strategy towards eliminating viral hepatitis[10,11]. The question arises whether SHM and the RIVM register the same cases of acute HCV infection among people with HIV in the Netherlands, and whether cases are missed by one registry or another. The aim of this study is to assess the completeness of these two national registries. In addition, we estimate the number of acute HCV infections among the HIV-positive population in the Netherlands, by means of capture-recapture analysis.

## Methods

### Data sources

Data were obtained from two national registries: the ATHENA national observational HIV cohort at SHM and the National Registry for Notifiable Diseases at the RIVM. We identified cases of acute HCV infection among adults (aged ≥18 years at HCV diagnosis) in the period of 2003-2016 in both registries. Both cases of a primary episode of acute HCV infection and reinfection were included; reinfections were treated as independent cases.

#### ATHENA cohort at SHM

Since 2000, SHM has managed the ATHENA cohort and is responsible for registering all HIV-positive people in care at the 26 HIV-treatment centres in the Netherlands[8]. At entry in HIV care, people are informed of the cohort and the purpose of data collection by their treating physician; participant consent follows an opt-out procedure (2% opt-out)[8]. After linkage to HIV care and registration at SHM, people are enrolled in the ATHENA cohort, which systematically collects demographic and clinical data from medical records, including information on HCV co-infection.

All cases of HCV infection were identified in the ATHENA cohort for the period of 2003 to 2016, based on the data lock in May 2017, followed by an update in January 2018. Diagnosis of HCV infection was based on available laboratory results of HCV-RNA and anti-HCV IgG[2]. *Acute HCV infection* definition corresponds to the case definitions by ECDC[12] and the NEAT preferred criteria [Grade A, Level II][13,14], and was defined as a positive anti-HCV IgG with a documented negative anti-HCV IgG within the previous 12 months, or a detectable HCV-RNA in the presence of either a documented negative HCV-RNA test or a documented anti-HCV IgG seroconversion within the previous 12 months. Additionally, cases of acute HCV reinfection were identified following sustained virological response (SVR), spontaneous clearance, or genotype-switch.

Additionally, cases of *chronic HCV infection* were defined as HCV-RNA positivity for >6 months. In case of an undetermined stage of HCV infection (i.e., acute or chronic), stage of infection was verified with the HIV-treating physician. Alternatively, if the stage of HCV infection remained unresolved, *other HCV cases* were included as a separate category. We defined the timing of HCV infection based on the date of the first lab diagnosis (RNA or antibody).

#### National Registry for Notifiable Diseases at RIVM

Since 1998, as a legal obligation enforced by the Dutch public health act (*Wet publieke gezondheid*)[15], treating physicians ànd diagnostic laboratories must report cases of HCV infection, together with demographic and epidemiological information, to the local Public Health Service (*Gemeentelijke of Gemeenschappelijke Gezondheidsdienst, GGD*). The GGD subsequently reports to the National Registry for Notifiable Diseases at the RIVM, using the web-based application OSIRIS[16]. As of 1 October 2003, only acute HCV infections need to be reported to the RIVM. The RIVM collects additional information on the HIV-serostatus of people with an acute HCV infection, and whether the HCV infection is a primary or reinfection, since 1 January 2013.

Cases of *acute HCV infection* were identified in the National Registry for Notifiable Diseases at the RIVM for the period of 2003 to 2016, based on the database as of 27 February 2017. The RIVM case definition corresponds to the SHM case definition (i.e. ECDC[12] and the NEAT preferred criteria [Grade A, Level II]) and, in addition, includes diagnoses based on a positive HCV-RNA associated with an elevated alanine aminotransferase according to the NEAT alternative criteria [Grade B, Level III][13,14]. At RIVM, cases by default were *acute HCV infections*, with either *confirmed HIV-positive* or *unknown HIV-serostatus* (captured since 2013). We determined the timing of HCV infection based on the reported date of lab diagnosis; if unavailable, the first date of illness, or the date of reporting to the RIVM was used.

### Case-linkage and statistical analysis

First, we described the annual number of reported cases by HCV/HIV transmission risk group and HIV serostatus for the period 2003-2016, using descriptive statistics.

Second, to facilitate capture-recapture analysis, cases of *acute HCV infection* were linked between the two databases for the period of 2013-2016. Cases were linked using a stepwise approach, based on: year and country of birth, sex, postal code (2 or 4 digits), municipal health service region, and date of HCV diagnosis (allowing a diagnostic timeframe). People who were living outside the Netherlands but who had been diagnosed and/or were in care in the Netherlands could not be matched on postal code and were matched based on municipal health service region. First, we linked the cases of acute HCV infection among cases with an *HIV-positive serostatus*. Second, we repeated the linkage and included additional cases of acute HCV infection reported to the RIVM with both *a confirmed positive* or *unknown HIV serostatus*.

Additionally, we assessed the influence of the distinction between acute and chronic HCV diagnosis on the registration in both databases. We linked RIVM cases of *acute HCV infection* among HIV-positive adults to *all* SHM cases (i.e., *acute, chronic*, and *other*). Subsequently, we repeated this analysis containing all RIVM cases of *acute HCV infection*, including cases with an *unknown HIV serostatus*.

#### The capture-recapture method

The capture-recapture method is derived from ecology and applied to epidemiological studies[17]. To estimate the number of cases in a population, cases are captured in one data source and then independently recaptured in a second data source. The capture-recapture method analyses the degree of overlap between the two data sources (**Figure 1**). The capture-recapture estimate is adequate when the following assumptions hold[18,19]: i) the study applies to a closed population, i.e., the definition or interpretation of the ‘target population’ is explicit; ii) linkage between the cases is possible and reliable; iii) the databases are functionally independent and do not rely on each other; vi) every case has the same probability to be included in each database.

**Figure 1.**
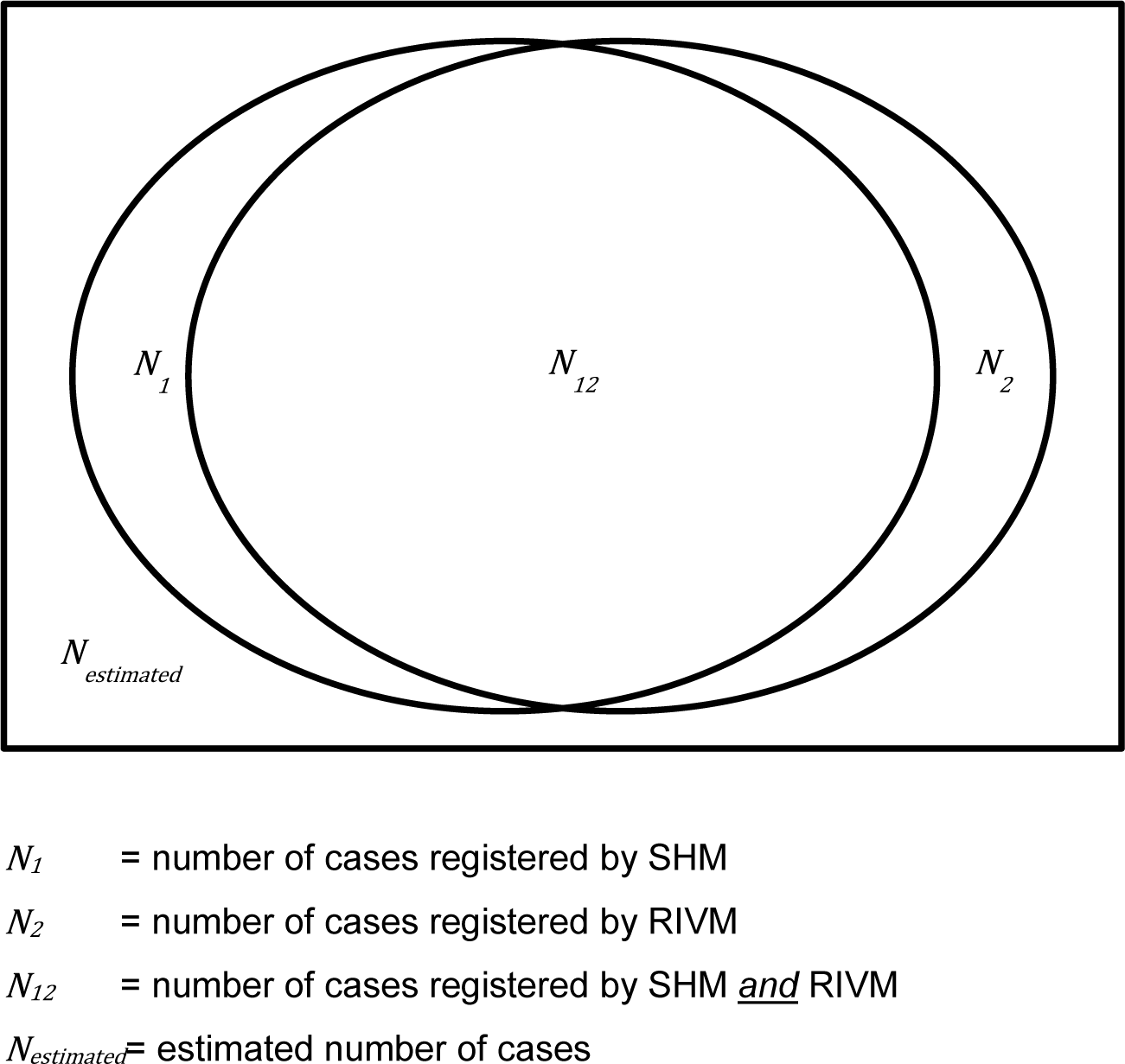
Venn diagram of capture-recapture analysis.

For this study, we performed a two-way capture-recapture analysis to estimate the total number of cases of acute HCV among HIV-positive people based on the two databases for the period of 2013 to 2016. We calculated the adjusted Petersen–Lincoln estimate[18,19] of the total number of cases in 2013-2016, and by calendar year (i.e., 2013, 2014, 2015 and 2016) separately:

Adjusted Petersen–Lincoln estimated number of cases:

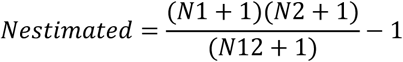

With the associated variance of the estimate of the total population:

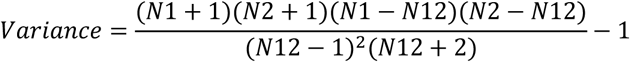

And the 95% confidence interval of the estimate of the total population:

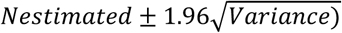

Where:

N_1_ = number of cases registered by SHM

N_2_ = number of cases registered by RIVM

N_12_ = number of cases registered by SHM *<u>and</u>* RIVM

N_estimated_ = estimated number of cases

### Ethics

At initiation, the ATHENA cohort[8] at SHM was approved by the institutional review board of all participating centres and patient consent is received by opting out. Data are pseudonymised to investigators and may be used for scientific purposes. The National Registry for Notifiable Diseases at RIVM anonymously registers mandatory communicable disease notifications[20], as a legal obligation enforced by the Dutch public health act[15] and International Health Regulations by the World Health Organisation[21].

For this study, we received approval from the steering committees of both institutions (i.e., RIVM and SHM). To secure non-tractability, personal identifiers were removed from the datasets before case-matching, using surrogate identifiers. A designated quality management coordinator safeguarded privacy protection and compliance with the European General Data Protection Regulation[22]. This work complies with the principles laid down in the Declaration of Helsinki[23].

## Results

### Acute HCV infections among adults in 2003-2016

Cases of HCV infection were extracted from both registries as described in **Figure 2**, and summarized in **Figure 3** and **Appendix table 1**.

**Figure 2.**
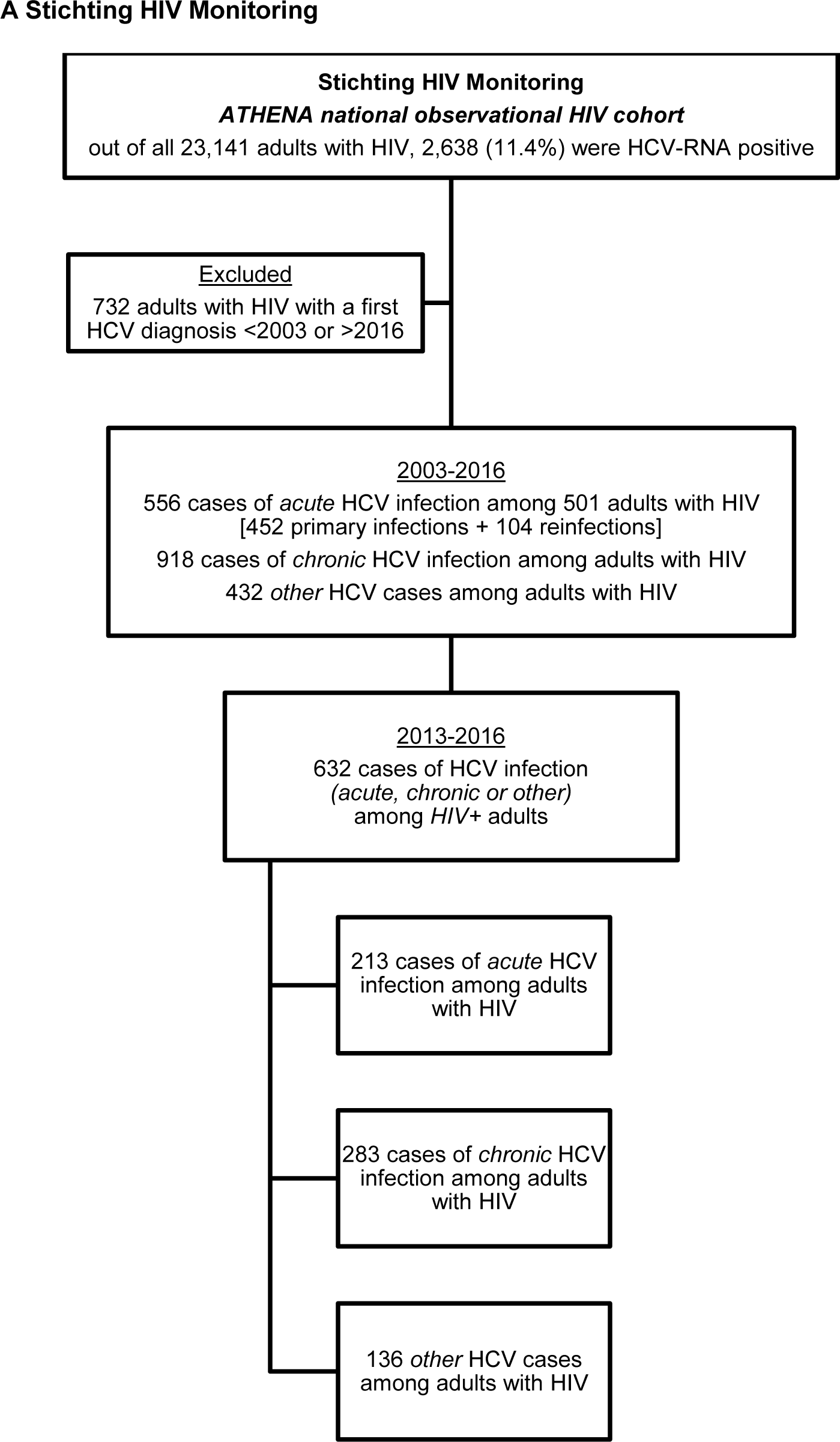
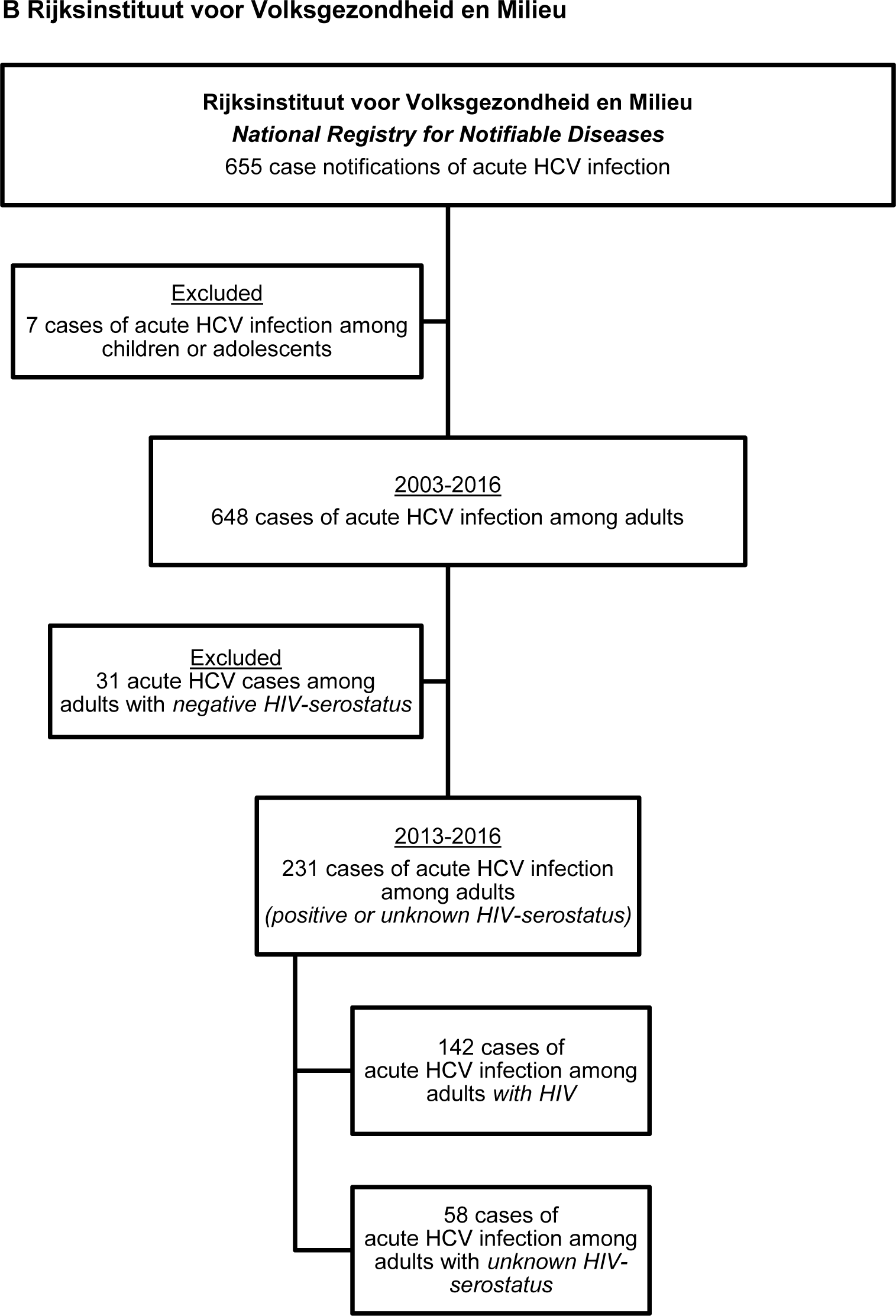
Acute hepatitis C case extraction from the registries at Stichting HIV Monitoring and the Rijksinstituut voor Volksgezondheid en Milieu.

**Figure 3.**
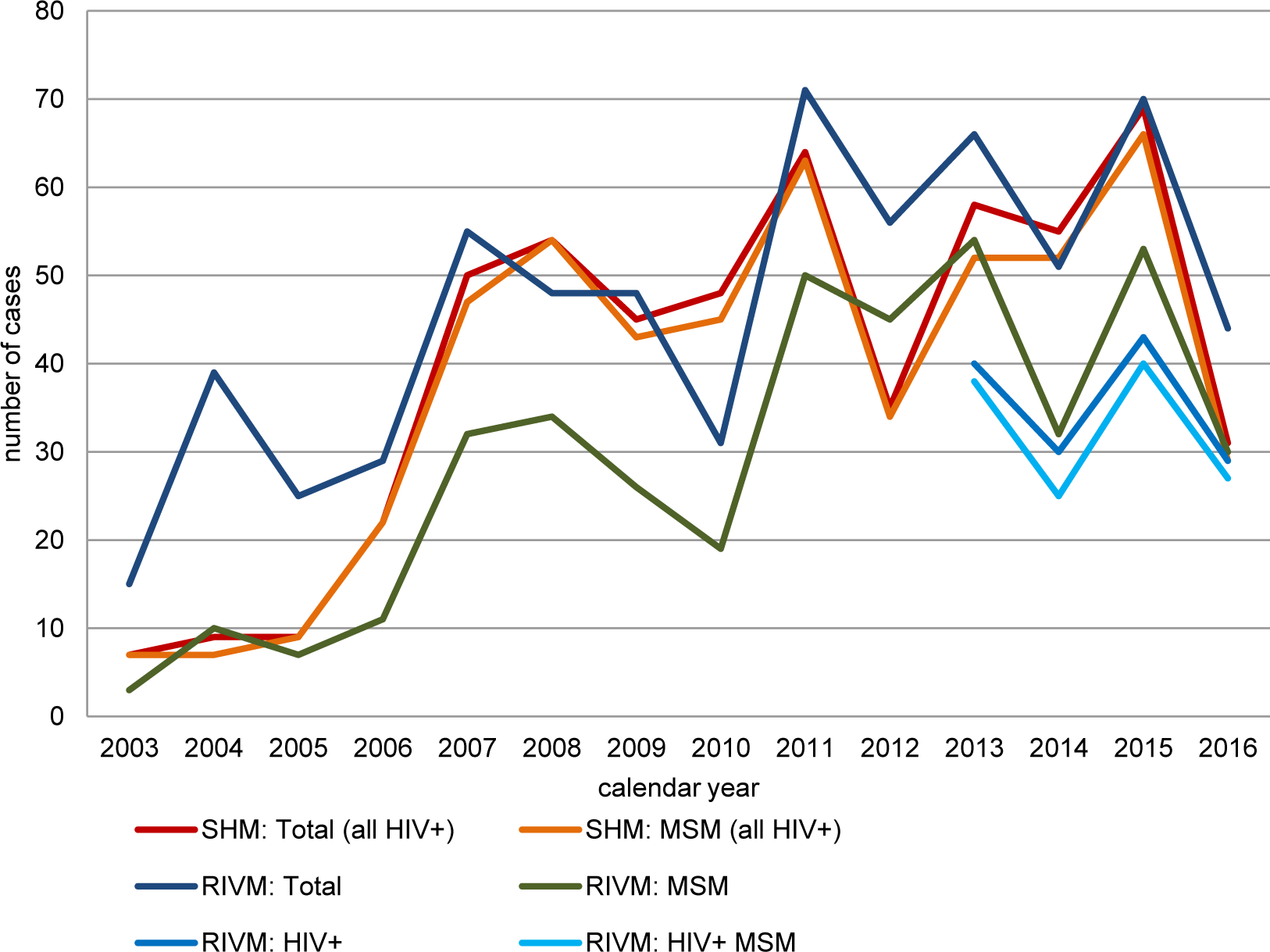
Number of acute hepatitis C infections, as registered by SHM and RIVM. Note: For SHM, all cases were among people with HIV. For RIVM, a positive HIV-serostatus is presented separately for men who have sex with men (MSM) only. In addition to the 131 MSM with HIV who acquired HCV through sexual contact, 15 cases of acute HCV infection were acquired through other sexual contact (N=3), injecting drug use (N=2), a needle stick- or bite-incident (N=1), or other/unknown routes (N=9). Also see appendix table 1.

#### ATHENA cohort at SHM

In total, 2,638 (11.4%) out of all 23,141 adults ever in HIV care and enrolled in ATHENA had a positive HCV-RNA test at least once (**Figure 2**). In the period of 2003-2016, 501 people were diagnosed with an acute primary HCV infection. In addition, 104 acute HCV reinfections were found among 86 people. The maximum number of acute HCV reinfections found per person was four. Altogether, we included 566 cases of *acute HCV infection* among 501 HIV-positive adults with HIV registered by SHM (**Appendix Table 1**).

We restricted our capture-recapture analysis to the 213 cases of *acute HCV infection* among 205 HIV-positive adults in the period 2013-2016 (**Figure 2, Table 1**). All but two cases were among males (N=211; 99.1%), the median age at HCV diagnosis was 43 years [IQR 36-50], and 171 (80.3%) were born in the Netherlands. Sexual contact among MSM was the main route of HIV infection (N=199; 93.4%). The remaining 12 cases were among people who had acquired HIV through heterosexual contact (N=6; 2.8%), injecting drug use (N=2; 0.9%), blood or blood products (N=2; 0.9%), or another or unknown route (N=4; 1.9%). Acute HCV was diagnosed a median 5.9 years [IQR 2.6-10.7] after HIV diagnosis.

**Table 1.**
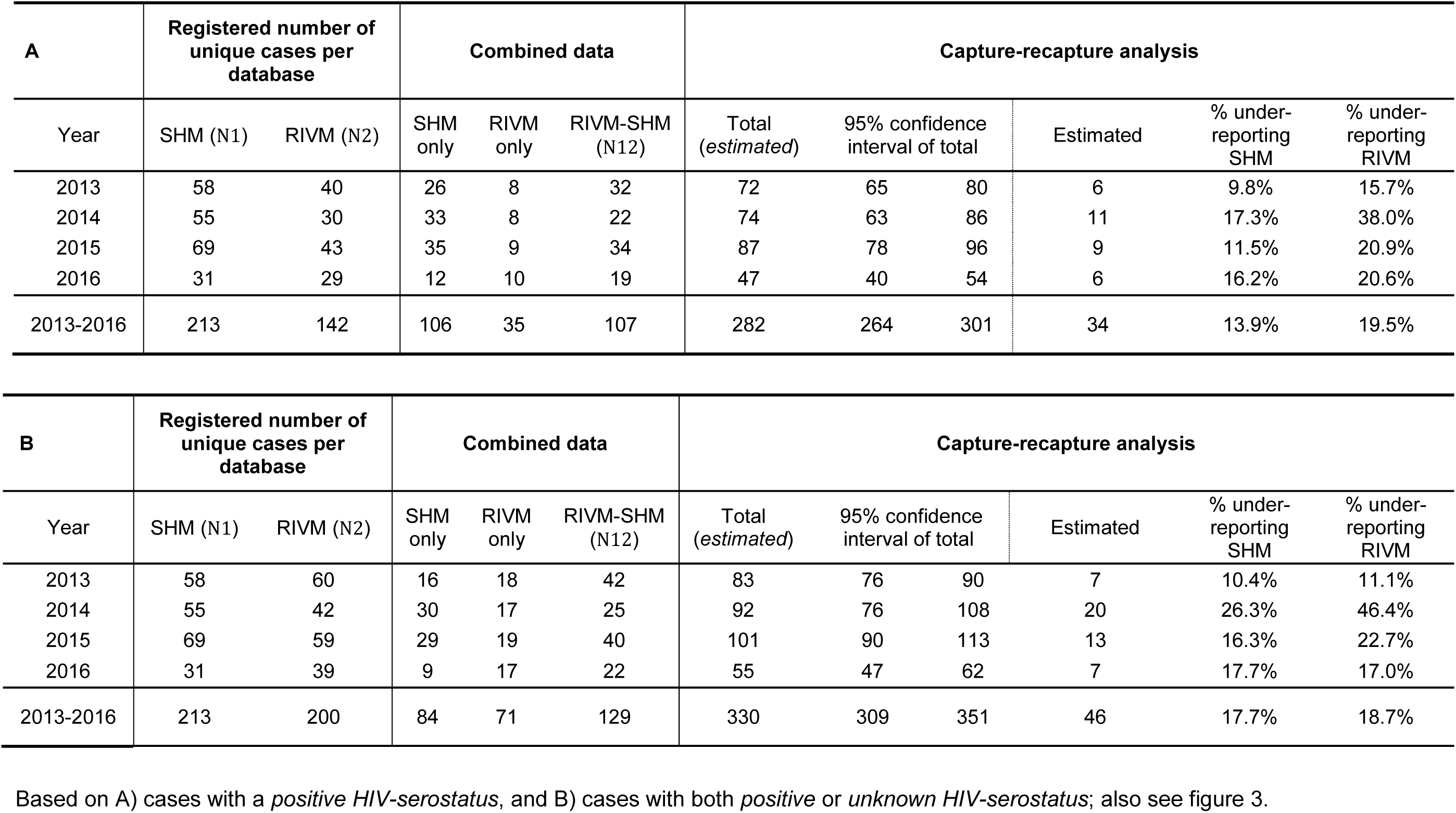
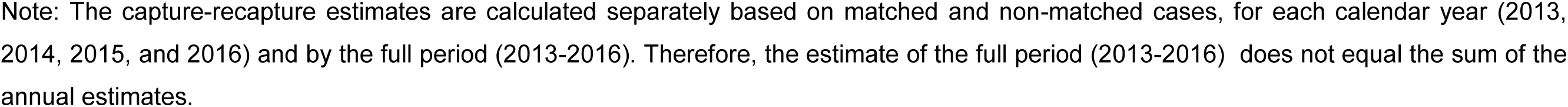
Number of acute hepatitis C virus infection cases: by data source, after case-linkage, and combined estimates based on capture-recapture analysis.

### Chronic HCV infections and other HCV cases

Additionally, 918 *chronic HCV infections* and 432 *other HCV cases* were registered in the period 2003-2016. Out of 432 *other HCV cases*, 357 people had a positive HCV antibody or RNA test followed by a negative HCV-RNA result, without having received HCV treatment. These people had likely spontaneously cleared the infection without treatment. Moreover, 75 out of 432 people were diagnosed with HCV based on a single HCV-RNA test result without further diagnostic information. In the period 2013-2016, 283 *chronic HCV infections* and 136 *other HCV cases* were registered. The median time between HIV and subsequent HCV diagnosis was 5.7 years [IQR 1.5-10.] and 5.6 years [IQR 0.5-11.9] for *chronic HCV infections* and *other HCV cases*, respectively.

#### National Registry for Notifiable Diseases at RIVM

In the period of 2003 to 2016, 655 cases of *acute HCV infection* were registered at the RIVM (**Figure 2**). Seven cases were excluded because they were among children or adolescents (<18 years of age); 5 out of 7 had acquired HCV through vertical transmission, leaving 648 cases of *acute HCV infection* among adults (**Appendix Table 1**). The median age at diagnosis was 43 years [IQR 35-49], and 89.0% were male (N=577). Two-thirds of the cases were reported among people who were born in the Netherlands (N=438; 67.6%). The main route of HCV transmission was sexual contact (N=434; 67.0%), followed by injecting drug use (N=50; 7.7%), or a needle stick or bite incident (N=16; 2.5%); the remaining 148 infections were acquired through another or unknown transmission route (22.8%). Overall, 406 out of 648 (62.7%) of all acute HCV infections were acquired through sex between men (i.e., MSM).

We restricted our capture-recapture analysis to the 231 cases of *acute HCV infection* in 2013-2016 (**Figure 2, Table 1**). Overall, 191 (82.7%) out of 231 HCV infections were known primary infections and 9 (3.9%) were reported as reinfections; data on primary/reinfection were unknown for the remaining 31 (13.4%) cases. In total, 142 out of 231 (61.5%) cases were reported among HIV-positive men, 58 (25.1%) had an unknown or missing HIV serostatus (28 of whom were MSM), and 31 (13.4%) were HIV-negative (11 of whom were MSM). Of note, 6 out of 9 reported HCV reinfections occurred in *HIV-positive* people; for the remaining 3 reinfections, the HIV-serostatus was unknown. No reinfections were reported among HIV-negative people.

All HIV-positive people were male, the median age at HCV diagnosis was 43 years [IQR 35-51], and 118 (83.1%) were born in the Netherlands. Sexual contact among MSM was the main route of HCV infection (N=130; 91.6%). The remaining 12 HCV infections were acquired through other sexual contact (N=2, heterosexual and unknown), injecting drug use (N=2), a needle stick or bite incident (N=1), or another or unknown route (N=7).

#### Capture-recapture analysis for 2013-2016

First, we linked the 213 and 142 cases of *acute HCV infection* among *HIV-positive* adults as registered in 2013-2016 by SHM and RIVM, respectively (**Table 1A**). In total, 107 cases were found in both registries: 50.2% of the 213 SHM cases could be linked to RIVM cases, and 75.4% of the 142 RIVM cases to SHM cases. The median time between diagnosis as documented by SHM and RIVM was 56 days [IQR 14-309]; 74 (69.2%) cases were documented by SHM first, 29 (27.1%) were documented by RIVM first, and 4 (3.7%) were diagnosed on the same date. Based on capture-recapture analysis, a total of 282 (95%CI: 264-301) acute HCV infections among HIV-positive adults were estimated for the period of 2013-2016. The estimated annual number of cases was 72 (95%CI: 65-80) in 2013, 74 (95%CI: 63-86) in 2014, 87 (95%CI: 78-96) in 2015 and 47 (95%CI: 40-54) in 2016 (**Figure 4A**). Based on these estimates, underreporting of acute HCV among HIV-positive people was 13.9% for SHM and 19.5% for RIVM.

**Figure 4.**
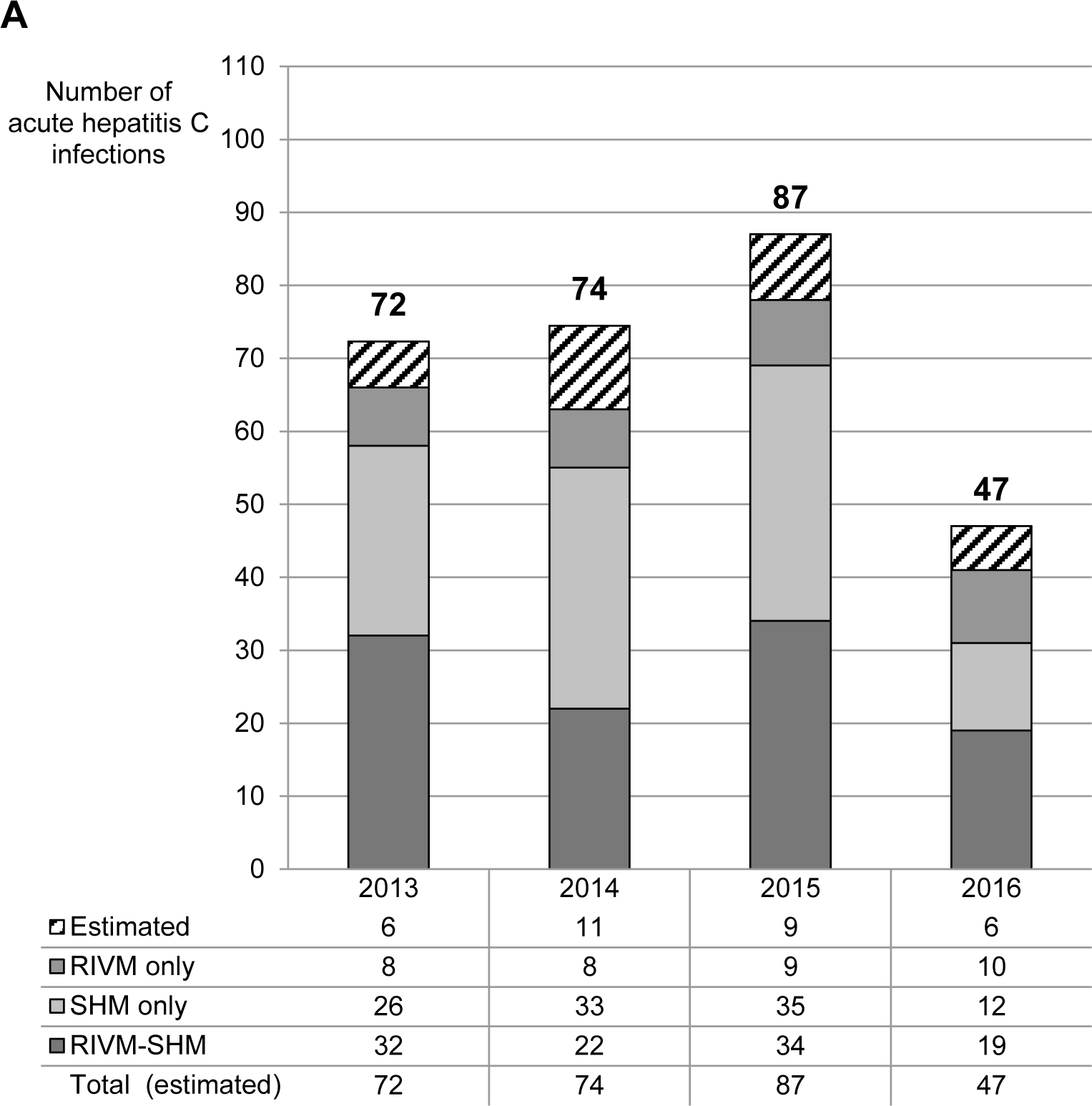
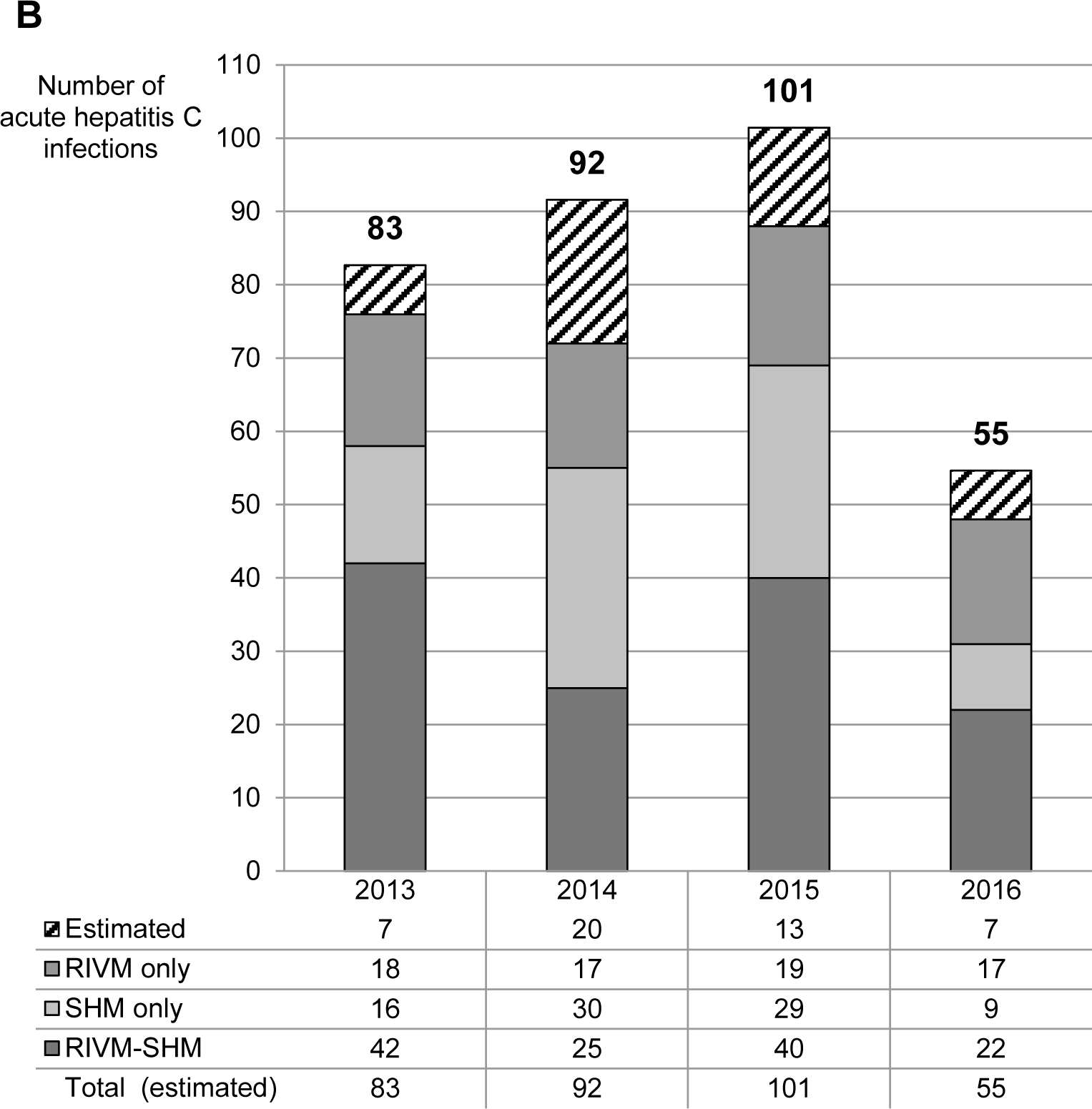
Acute hepatitis C infections among adults with HIV in the Netherlands: capture-recapture estimates. Based on A) cases with a positive HIV-serostatus, and B) cases with both positive or unknown HIV-serostatus; also see table 1.

Second, the addition of 58 RIVM-cases with an *unknown HIV-serostatus* led to an increase in the number of matches (**Table 1B**). In total, 129 cases were found in both registries: 60.6% of the 213 SHM-cases, and 64.5% of the 200 RIVM cases. The median time between diagnosis as documented by SHM and RIVM was 56 days [IQR 14-341]; 90 (69.8%) cases were documented by SHM first, 36 (27.9%) were documented by RIVM first, and 3 (2.3%) were diagnosed on the same date. The addition of cases with an *unknown HIV serostatus* led to an increase of the overall estimated total number of acute HCV infections among HIV-positive adults: from 282 (95%CI: 264-301) to 330 (95%CI: 309-351) estimated cases in the period of 2013-2016 (**Figure 4B**). With the addition of RIVM cases with an unknown HIV serostatus, underreporting of acute HCV among HIV-positive people was 17.7% for SHM and 18.7% for RIVM.

#### Chronic HCV infections and other HCV cases

Additionally, we linked *all* 632 cases of HCV infection among HIV positive adults registered by SHM (i.e., *acute, chronic*, and *other*), with the 142 cases of *acute HCV infection* adults who were *HIV positive*, registered by the RIVM (**Appendix Table 2**). All but two RIVM-cases could be linked to an acute, chronic, or *other HCV case* registered by SHM (98.6%; 140 out of 142). Furthermore, when we added the 58 RIVM-cases of *acute HCV infection* among people with an *unknown HIV serostatus* -- i.e. when we linked the 632 SHM cases with the 200 RIVM cases -- we could link 175 cases of HCV infection, indicating underreporting of HIV-positivity at RIVM and missed acute cases in the SHM data.

## Discussion

Based on data from the two national registries (SHM and RIVM), we estimated 282-330 cases of acute HCV infection among HIV-positive adults in the Netherlands, for the period of 2013-2016. Underreporting was estimated at 14-18% for SHM and 19-20% for RIVM. Underreporting could partially be explained by an unknown HIV-serostatus at RIVM, or by differences between RIVM and SHM in HCV stage at first diagnosis (acute or chronic). From 2003-2016, the predominant route of HCV transmission was sexual transmission among (HIV-positive) MSM.

The results should be interpreted while taking into account the study’s limitations. Notably, capture-recapture analysis is based on several assumptions, which influence the accuracy and reliability of the presented estimates[18,19]. Our study fulfils all four assumptions. However, incorrect case linkage and non-linkage cannot be ruled out because linkage was based on personal characteristics, and not on personal identifiers.

Our annual estimates of the number of acute HCV infections among people with HIV are in line with HCV incidence studies performed among HIV-positive MSM both in Amsterdam[24] and across Europe[25]. The estimated number of acute HCV infections was lower in 2016 than in to 2013, 2014 and 2015, which could be explained by a delay in registration, a change in the frequency of HCV testing[26], or an actual decrease in HCV incidence among HIV-positive people in the Netherlands, most likely due to increased DAA uptake. The latter is in line with recently published findings covering 17 out of 26 Dutch HIV-treatment centres, relating the high DAA uptake to a lower HCV incidence among HIV-positive MSM in 2016, than in to 2014[5]. Of note, annual numbers are known to fluctuate over time, also before 2013, and recent numbers reported by the RIVM showed a subsequent increase in the total number of HCV infections for 2017 compared to 2016[4].

Underreporting at both registries could be explained by several factors. At SHM, underreporting of acute HCV infections could be due to missed or incomplete screening for HCV by the HIV treating physician. Virtually all RIVM cases of acute HCV infection could be identified at SHM, although some RIVM cases of acute HCV were registered as chronic HCV at SHM. Previous analyses of the SHM have data shown substantial variation in routine HCV screening for all HIV-positive people at entry into HIV care, as well as in annual HCV screening among risk groups (i.e., MSM)[2]. This may, to some extent, be explained by centres applying a policy of targeted screening guided by the presence of incident transaminase elevations[27]. Furthermore, complimentary qualitative investigation has identified additional guideline, patient, physician, and environmental factors influencing the compliance to guidelines for laboratory monitoring in outpatient HIV care in the Netherlands[28]. At the RIVM, cases are reported through the local GGD, which receives case notifications from the diagnostic laboratories and physicians in or outside of HIV care. A marked reduction in HCV screening at some Dutch STI clinics among HIV-positive MSM could explain missed diagnoses and, as such, underreporting of acute HCV infections, specifically among HIV-positive people not engaged in HIV care or not screened for HCV in HIV care. This is largely an effect of a change in screening policy by the Amsterdam STI clinic, the largest in the Netherlands, which had to discontinue HCV screening due to financial constraints in 2014; although HCV screening in HIV-positive MSM was restarted in 2017. However, the acute HCV infections among HIV positive people should have been diagnosed in HIV care and reported to the RIVM. Additionally, a large proportion of RIVM cases had a missing or unknown HIV-serostatus. Unknown HIV serostatus could be due to a notification by the laboratory (that is unaware of HIV-serostatus), or due to lack of HIV testing among HCV-positive people. The relatively high HCV prevalence observed among (HIV-negative) MSM starting PrEP[29] underscores the need for routine combined STI/HCV screening among MSM at high risk for HIV infection.

Our study confirmed that linkage of two national databases is possible, and can provide reliable surveillance data with additional value. Previously, van Leth *et al*., linked the national tuberculosis registry to SHM to assess the TB-HIV prevalence in the Netherlands[30]. Data linkage studies are versatile and can strengthen multi-disease surveillance. Infectious disease surveillance data are essential for the planning and evaluation of public health activities. The sensitivity of surveillance systems relies on the likelihood of the infections being identified, diagnosed, and reported. We therefore recommend that all HCV infections, both acute and chronic, are registered at the National Registry for Notifiable Diseases at the RIVM, as in practice the timing of diagnosis and reporting by different health care workers can differ. Limiting the notification and registration to acute HCV infections only was primarily motivated by the need to trace sources and contacts. However, recent national action plan to eliminate hepatitis requires comprehensive detection and registration of all HCV infections[10,11].

To control and eventually eliminate HCV from the HIV-positive population in the Netherlands and beyond, all HIV/HCV co-infected people should be diagnosed, linked to, and retained in care[10,11]. Registration is essential to monitor trends in the epidemic, and to assess the impact of treatment and other interventions on HCV incidence. The high rate of HCV reinfections among HIV-positive MSM[2,31] warrants the need for increased awareness and prevention measures for high risk MSM, as well as structural repeat screening of HCV infection[32,33]. We call for increased attention for both documentation of HCV cases in medical records and disease notification at RIVM, by all health professionals. Surveillance data should ideally include both acute and chronic HCV infections, and be able to distinguish between acute and chronic infections, and initial and reinfections. Robust HCV surveillance is essential to monitor the impact of prevention programs and DAA treatment uptake – on both the individual and public health level.

## Data Availability

The data used in this study are confidential and would not be shared. The ATHENA cohort data are available on request at SHM; for more information: https://www.hiv-monitoring.nl/english/research/research-projects/research-proposal/.

